# Development of a tool to prioritize the monitoring of COVID-19 patients by public health teams

**DOI:** 10.1101/2021.04.08.21254922

**Authors:** Andres I. Vecino-Ortiz, Nicolás Guzman-Tordecilla, Yenny Fernanda Guzmán Ruiz, Rolando Enrique Peñaloza-Quintero, Julián A. Fernández-Niño, Fernando Ruiz Gomez, Antonio J. Trujillo

## Abstract

**Background:** In the context of the COVID-19 pandemic, public health teams have struggled to conduct monitoring for confirmed or suspicious COVID-19 patients. However, monitoring these patients is critical to improving the chances of survival, and therefore, a prioritization strategy for these patients is warranted. This study developed a monitoring algorithm for COVID-19 patients for the Colombian Ministry of Health and Social Protection (MOH).

**Methods:** This work included 1) a literature review, 2) consultations with MOH and National Institute of Health officials, and 3) data analysis of all positive COVID-19 cases and their outcomes. We used clinical and socioeconomic variables to develop a set of risk categories to identify severe cases of COVID-19.

**Results:** This tool provided four different risk categories for COVID-19 patients. As soon as the time of diagnosis, this tool can identify 91% of all severe and fatal COVID-19 cases within the first two risk categories.

**Conclusion:** This tool is a low-cost strategy to prioritize patients at higher risk of experiencing severe COVID-19. This tool was developed so public health teams can focus their scarce monitoring resources on individuals at higher mortality risk. This tool can be easily adapted to the context of other lower and middle-income countries. Policymakers would benefit from this low-cost strategy to reduce COVID-19 mortality, particularly during outbreaks.

## INTRODUCTION

The novel coronavirus SARS-CoV-2 (COVID-19) spread across the world and placed many health systems under unprecedented strain. During the first months of the pandemic, a rush to understand early treatment options for COVID-19 was made. Towards the midst of 2020, there was a relatively good understanding of early strategies to reduce the disease’s lethality to complement and improve the efficacy of more traditional public health interventions as social distance, lockdowns, hand washing, and contact tracing ^1,2^. However, overwhelmed health systems struggled to provide appropriate monitoring to all identified cases, particularly during outbreaks. Moreover, monitoring is challenging because it requires high upfront investments in infrastructure, planning, trained personnel, and information systems. Despite the high positive externalities, monitoring COVID-19 confronts organizational barriers that slow down its implementation.

The severity of COVID-19 infection is heterogeneous, and the fatality rate in the general population is estimated around 1%^3^. Some individuals are asymptomatic or experience mild flu-like symptoms, but others suffer a severe illness that can lead to death. The literature shows a higher probability of severe disease and deaths among older individuals with comorbidities such as diabetes, cardiovascular diseases, obesity, and respiratory diseases ^4^. COVID-19 cases with these conditions benefit from early and constant monitoring to seek advanced health care as soon as possible. However, given the limited resources available to monitor individuals, prioritizing severe disease risk categories is an urgent need. Such need is particularly true during outbreaks when the pressure on health systems is higher. Especially during high-demand times, it is necessary to identify early those with the highest probability of complications to receive faster life-saving care.

In this study, we developed and tested a strategy to prioritize monitoring systems for COVID-19 patients by directing monitoring resources to those in need. The study describes the development of this tool. This tool can be of use by other lower and middle-income countries (LMIC). This tool will also benefit other countries by reducing negative externalities towards other health care services.

## METHODS

In this work, we first developed a literature review to identify relevant variables for the prioritization tool. Based on the predictors identified during the review, we conducted the preliminary design of the prioritization tool in consultation with officials from the Colombian Ministry of Health and Social Protection and the National Institute of Health of Colombia (MH-NIH). Finally, we conducted the data analysis to assess the validity of the prioritization algorithm.

### Literature review

The literature review aimed to identify effect sizes for variables that predict a higher likelihood of severe or fatal COVID-19. We conducted a MEDLINE and Google Scholar review of papers in English and Spanish, in both preprint and peer-reviewed journals, and published before October 1, 2020. The following search terms were used in English: *Novel Coronavirus, Epidemic, Pandemics, Sars-Cov-2, COVID-19, Coronavirus Infections, Complications, Mortality, Intensive Care Unit, Disease Severity, Mortality rate, Fatality rate, Deaths, Severity, Obesity, Body Mass Index, Total Body Weight, Smoking, Comorbidity, Cerebrovascular, Meta-Analysis*.

In Spanish, the terms were: Nuevo coronavirus, Epidemia, Pandemia, *Sars-Cov-2, COVID-19, Infección por* coronavirus, Complicaciones, Mortalidad, Unidad de Cuidado intensivo, UCI, *severidad de la enfermedad, Tasa de mortalidad, Tasa de fatalidad, Muerte, Severidad, Obesidad, Índice de masa corporal, Exceso de peso, Sobrepeso, Fumar, Comorbilidad, Enfermedad Cerebrovascular, Meta-análisis*.

*Moreover*, we identified the papers cited in the reference list to widen the search.

The inclusion criteria for the selection of papers were:

1. Papers describing severe or fatal COVID-19 (defined as a patient requiring admission to an intensive care unit, intubation, or dying from COVID-19).
2. Papers including risk and predictor factors that have been measured empirically or have been described as high-risk factor for severe or fatal COVID-19.

The exclusion criteria for the selection of papers were:

1. Papers were abstracts, conference proceedings, or book chapters
2. Papers that did not directly address risk factors for severe or fatal COVID-19.
3. We did not exclude papers based on geography, gender, or age.

Our initial search identified 8,319 papers. Excluding 1,081 duplicated manuscripts, our final search identified 7,238 papers. Two reviewers screened titles and abstracts yielding 194 papers for full-text review, and selected 12 papers for inclusion. The search strategy can be seen in figure 1. In these papers, we obtained a list of all potential variables with more predictive value.

**Figure 1.**
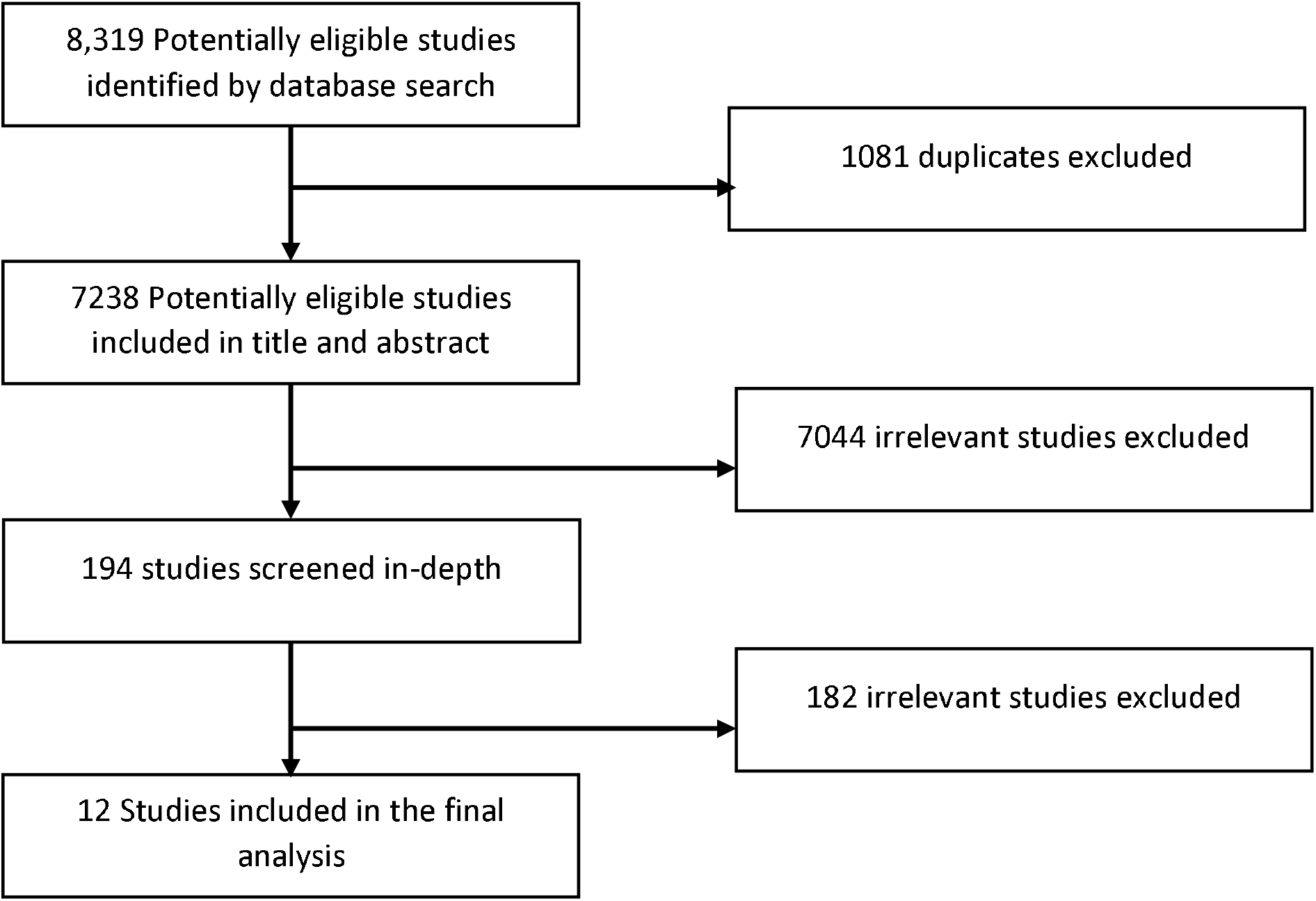
Review of Literature and Study Selection.

### Data sources

We used official combined data for all 1) confirmed, 2) suspected, and 3) negative Coronavirus cases in Colombia in the general population (excludes specific closed populations including military, police personnel, and institutionalized individuals, which represent a small percentage of the population) through the Ministry of Health (MOH) and National Health Institute (NIH) platforms (SEGCOVID19 and SIVIGILA, respectively), between March 6 and October 11, 2020. This dataset ascertains data on all individuals’ health, demographic, and socioeconomic characteristics with a confirmed, possible or negative diagnosis of COVID-19^5^.

SEGCOVID19 is the main platform where all the data from the national contact tracing program (called PRASS, the Spanish acronym for Sustainable Program for Tests, Tracing, and Selective Isolation) is collected and consolidated. This system collects information from both notifications issued by the National Health Institute system (SIVIGILA) and cases and contacts identified by insurers. The system is critical to trace transmission chains and identify suspected contacts. SEGCOVID19 also collects information from all tests performed in the country and gathers data from databases recording health care service provision, civil registration, and vital statistics records. We used both records to ensure that we captured as many variables as possible from every single case so that the prediction model would be more comprehensive.

### Selection of variables

We used the following criteria to select variables for the algorithm:

1. A large effect size of the variable predicting lethality by COVID-19
2. A reliable effect size of the variable predicting lethality by COVID-19
3. The availability of the variable in the data obtained from SEGCOVID19 and SIVIGILA.

The variables selected as per these selection criteria are described below.

1. *Comorbidities:* This variable includes those who reported having at least one of the following diseases or conditions: Cancer, HIV, cerebrovascular disease, high blood pressure (HBP), heart disease, immunosuppressed, cardiovascular disease, diabetes mellitus, smoking, obesity, and kidney disease. It should be noted that the dataset does not provide information about whether these diseases were controlled. Individuals with missing data on comorbidities were assumed to have no comorbidities.
2. *Age*: Measured in years old that includes both men and women.
3. *Socioeconomic status:* The *socioeconomic status* (SES) is a variable with six categories commonly used by the national government to target subsidies, and it has been treated as a proxy for socioeconomic status in previous research in Colombia^6^. Observations in stratum 1 to 3 were categorized as low SES and those between 4 to 6 as medium-high SES^7^.
4. *Pregnancy*: Women who are pregnant, regardless of the stage of their pregnancy.

The outcome variable (severe COVID-19) is defined as an individual being admitted to intermediate care/intensive care or dying from COVID-19.

### Preliminary design and consultation with the MH-NIH

After the literature review, we held three meetings to consult with government officials on the feasibility of using these variables. Considering current data systems, we designed a preliminary prioritization algorithm with four different risk categories to classify individuals who are more likely to experience severe disease or death. As monitoring services, experience changes in capacity, training, or during outbreaks, the co-designed algorithm provides a hierarchical pathway to prioritize case monitoring flexible to changes in its supply or demand. Hence, monitoring personnel can focus on cases that have a higher chance of becoming severe. This study was approved by the Institutional Review Board of the Johns Hopkins Bloomberg School of Public Health and deemed not human subjects research (IRB number: 14144).

### Data analysis

We conducted a univariate analysis of the variables selected. Further and based on the consultations, we assessed the conditional probabilities of detecting a severe COVID-19 case for each of the four risk categories. Operational characteristics were observed using Receiver Operating Characteristic (ROC) curves and based on these, adjustments on the order of the variables were made.

The operational characteristics obtained from the algorithm were calculated under the following assumptions:

1. The data obtained for the analysis are assumed to be a complete set of all severe and fatal cases.
2. All cases that did not report information on the severity of illness were considered non-severe cases.

## RESULTS

### Literature review and consultations with the MH-NIH

Based on the literature review, a prioritization of the most relevabt variables to predict severe COVID-19 was carried out. This process included the identification of variables according to severity (Table 1).

**Table 1.**
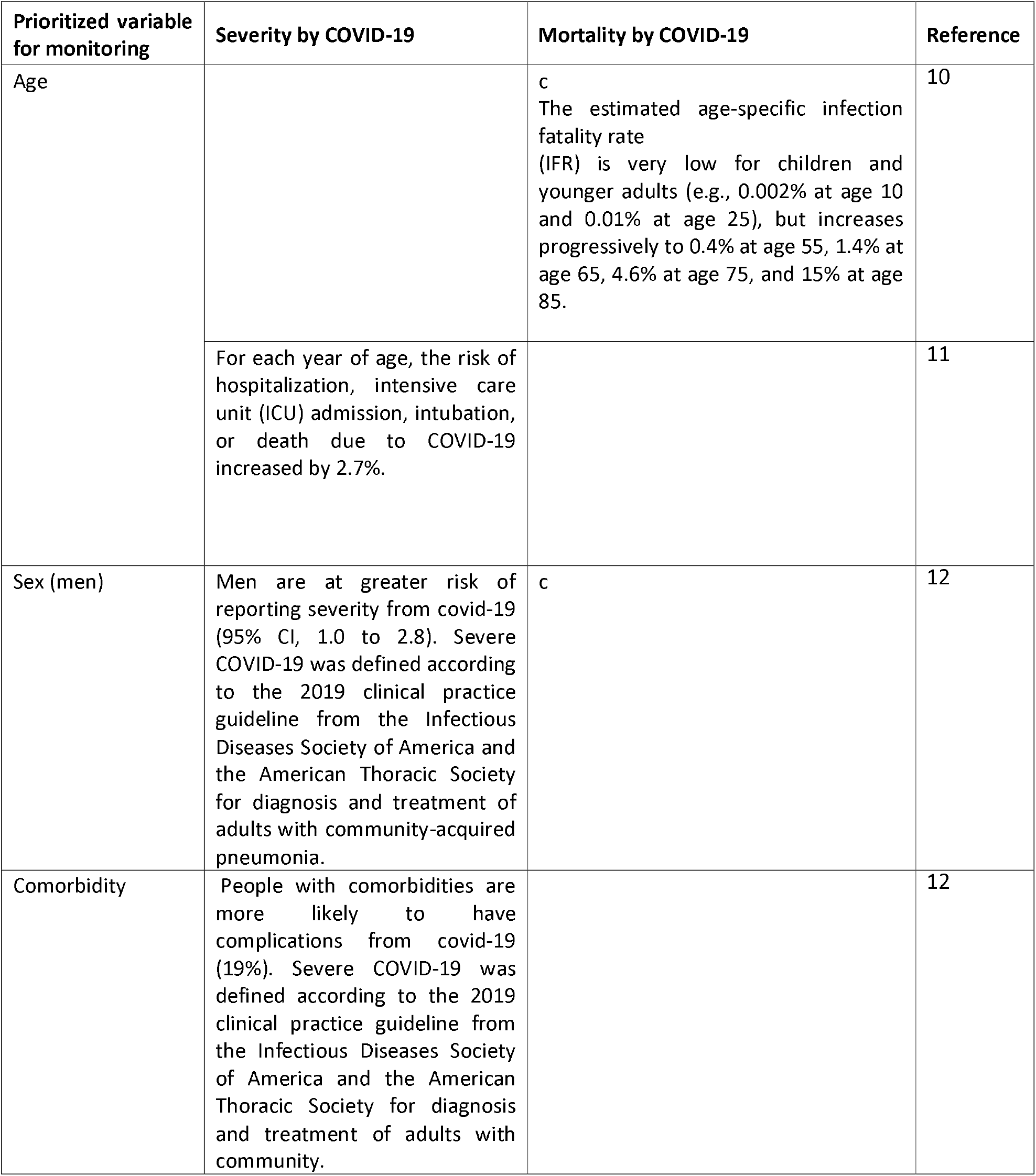
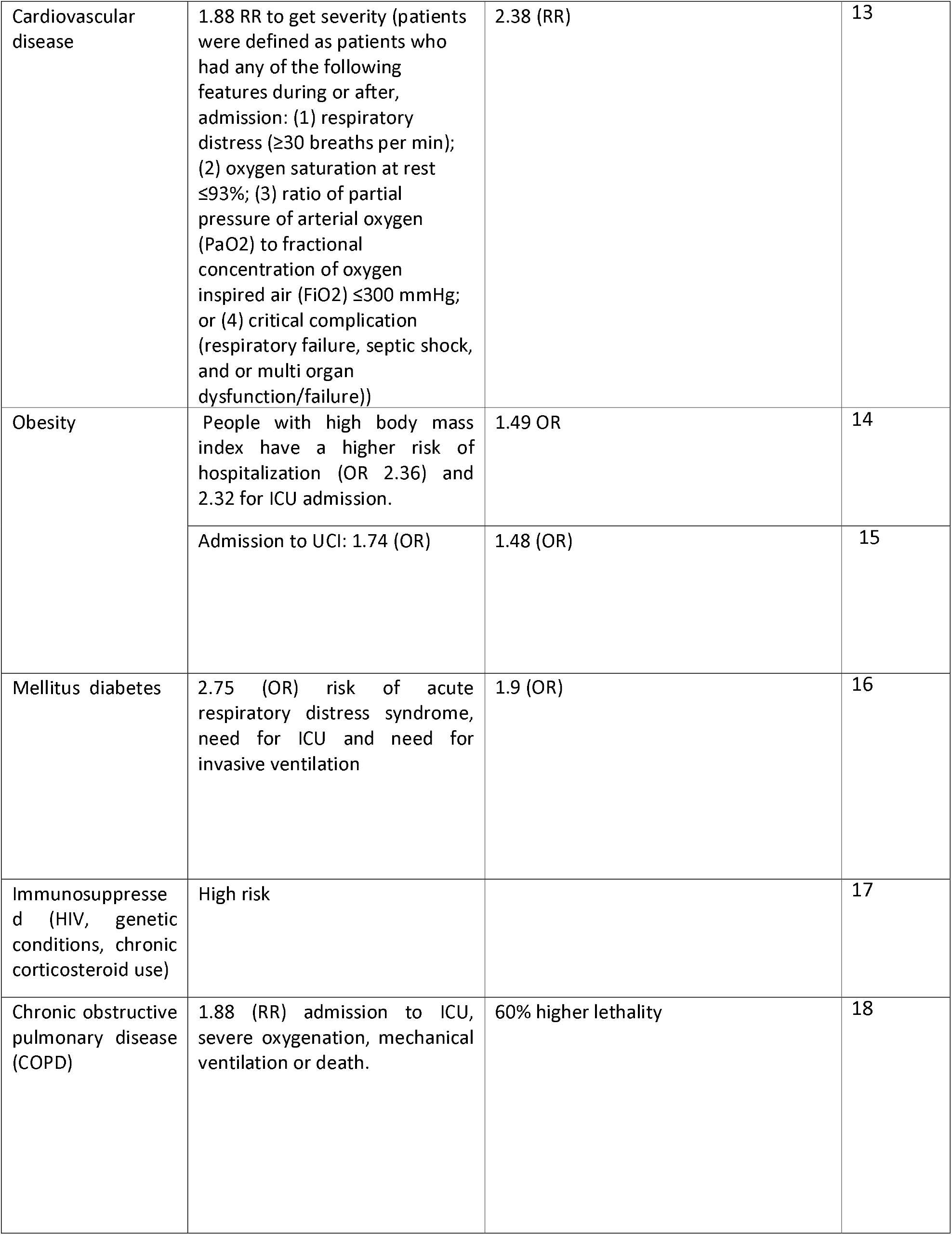
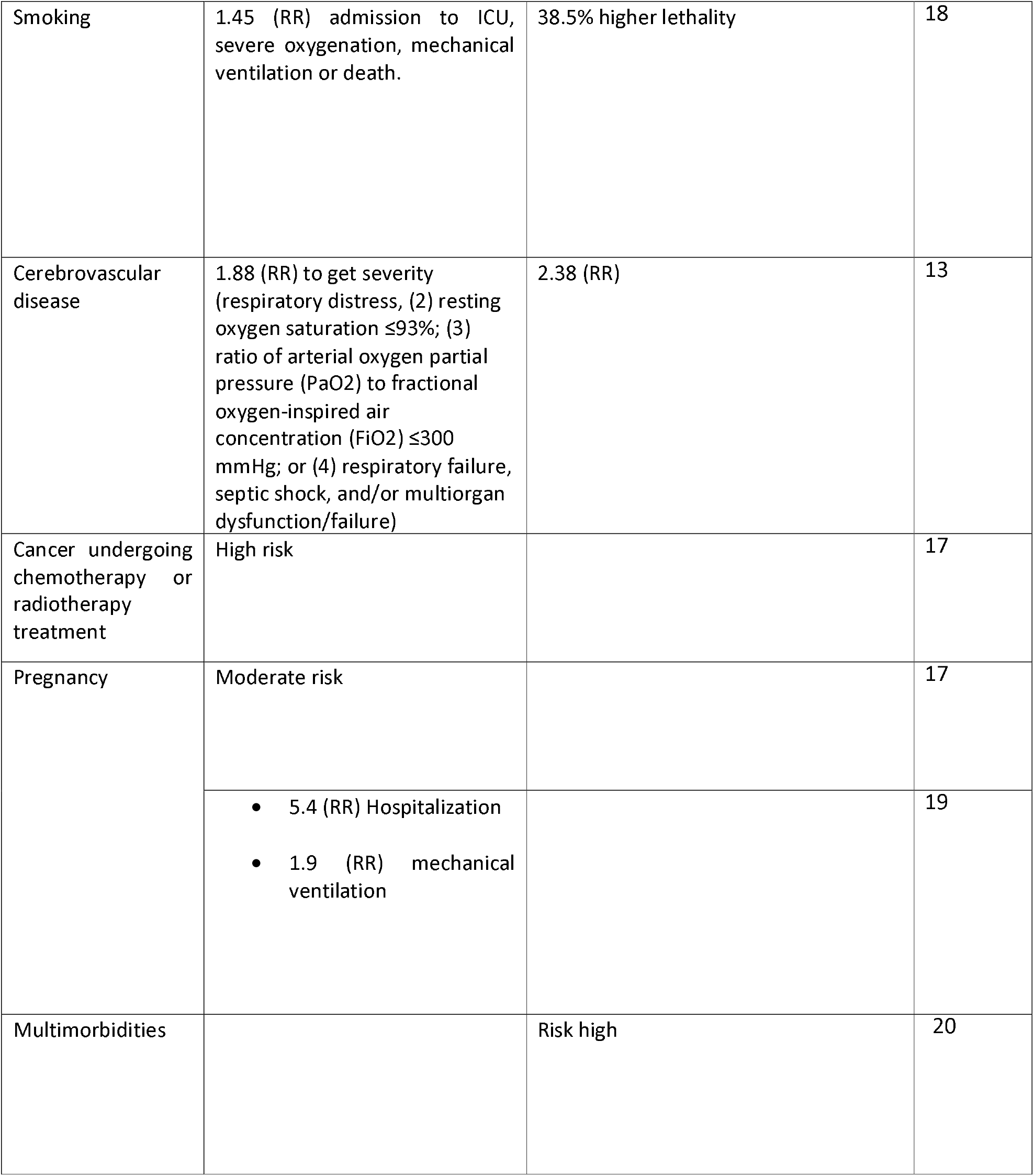
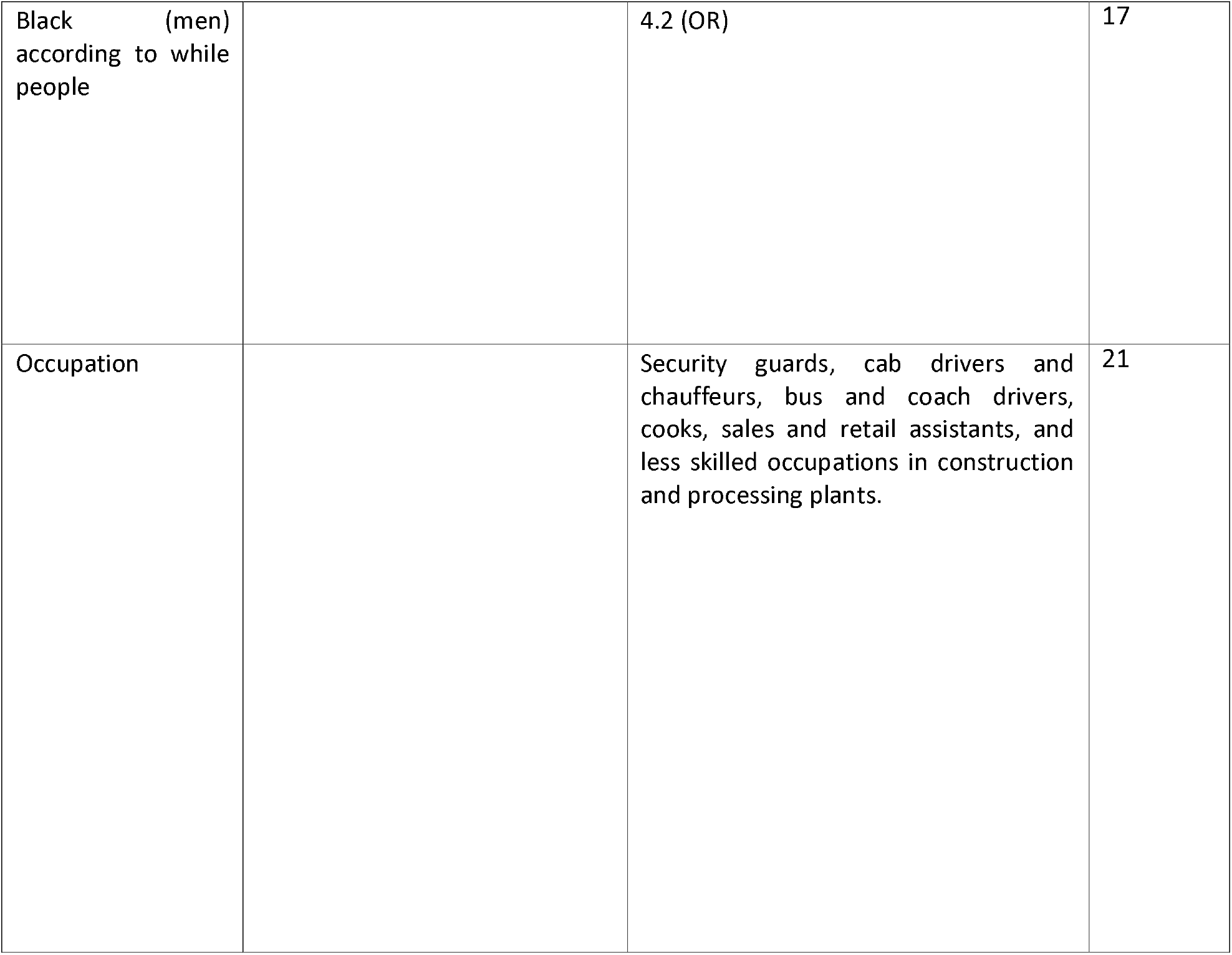
Case Monitoring Literature Review Matrix.

| Prioritized variable for monitoring | Severity by COVID-19 | Mortality by COVID-19 | Reference |
| --- | --- | --- | --- |
| Age |  | c<br>The estimated age-specific infection fatality rate (IFR) is very low for children and younger adults (e.g., 0.002% at age 10 and 0.01% at age 25), but increases progressively to 0.4% at age 55, 1.4% at age 65, 4.6% at age 75, and 15% at age 85. | 10 |
|  | For each year of age, the risk of hospitalization, intensive care unit (ICU) admission, intubation, or death due to COVID-19 increased by 2.7%. |  | 11 |
| Sex (men) | Men are at greater risk of reporting severity from covid-19 (95% CI, 1.0 to 2.8). Severe COVID-19 was defined according to the 2019 clinical practice guideline from the Infectious Diseases Society of America and the American Thoracic Society for diagnosis and treatment of adults with community-acquired pneumonia. | c | 12 |
| Comorbidity | People with comorbidities are more likely to have complications from covid-19 (19%). Severe COVID-19 was defined according to the 2019 clinical practice guideline from the Infectious Diseases Society of America and the American Thoracic Society for diagnosis and treatment of adults with community. |  | 12 |
| Cardiovascular disease | 1.88 RR to get severity (patients were defined as patients who had any of the following features during or after, admission: (1) respiratory distress ($\geq 30$ breaths per min); (2) oxygen saturation at rest $\leq 93\%$; (3) ratio of partial pressure of arterial oxygen (PaO<sub>2</sub>) to fractional concentration of oxygen inspired air (FiO<sub>2</sub>) $\leq 300$ mmHg; or (4) critical complication (respiratory failure, septic shock, and or multi organ dysfunction/failure)) | 2.38 (RR) | 13 |
| Obesity | People with high body mass index have a higher risk of hospitalization (OR 2.36) and 2.32 for ICU admission. | 1.49 OR | 14 |
|  | Admission to UCI: 1.74 (OR) | 1.48 (OR) | 15 |
| Mellitus diabetes | 2.75 (OR) risk of acute respiratory distress syndrome, need for ICU and need for invasive ventilation | 1.9 (OR) | 16 |
| Immunosuppressed (HIV, genetic conditions, chronic corticosteroid use) | High risk |  | 17 |
| Chronic obstructive pulmonary disease (COPD) | 1.88 (RR) admission to ICU, severe oxygenation, mechanical ventilation or death. | 60% higher lethality | 18 |
| Smoking | 1.45 (RR) admission to ICU, severe oxygenation, mechanical ventilation or death. | 38.5% higher lethality | 18 |
| Cerebrovascular disease | 1.88 (RR) to get severity (respiratory distress, (2) resting oxygen saturation $\leq 93\%$; (3) ratio of arterial oxygen partial pressure (PaO<sub>2</sub>) to fractional oxygen-inspired air concentration (FiO<sub>2</sub>) $\leq 300$ mmHg; or (4) respiratory failure, septic shock, and/or multiorgan dysfunction/failure) | 2.38 (RR) | 13 |
| Cancer undergoing chemotherapy or radiotherapy treatment | High risk |  | 17 |
| Pregnancy | Moderate risk |  | 17 |
|  | • 5.4 (RR) Hospitalization<br>• 1.9 (RR) mechanical ventilation |  | 19 |
| Multimorbidities |  | Risk high | 20 |
| Black (men)<br>according to white<br>people |  | 4.2 (OR) | 17 |
| Occupation |  | Security guards, cab drivers and chauffeurs, bus and coach drivers, cooks, sales and retail assistants, and less skilled occupations in construction and processing plants. | 21 |

Some variables identified in the literature as relevant were not included in the final algorithm because they were either not available in the database (e.g., cancer undergoing chemotherapy or radiotherapy treatment) or its inclusion was logistically challenging (e.g., occupation). From these consultations, adjustments were made to the algorithms according to logistical feasibility and the available data. For this reason, as the measurement capabilities of these variables are improved, they might be considered in new versions of this algorithm. This algorithm is developed for the general population. Special populations (e.g., military, prison populations) or other closed populations may take elements of this algorithm. However, the data with which this algorithm was evaluated are not representative of those populations.

### Data analysis

#### Descriptive

The total sample size was comprised of 972,059 cases; 51% were women. The mean age for the total sample was 38.90 years old (40-59 years: 29,2%, 60 and more years: 15%), and 94% of the cases were considered low SES. Also, 9% of the cases had *comorbidities*. 2.4% of the cases presented severe COVID-19 (table 2). It should be noted that the distribution of cases (particularly mild and asymptomatic cases) does not necessarily correspond to the distribution of past infection (seroprevalence).

**Table 2.** Descriptive analysis of the variables included in the monitoring algorithm.

|  |  |
| --- | --- |
| **Variable** | n= 972,059 (%) |
| **Age (years old)** |  |
| Mean (standard deviation) | 39 (18.86) |
| Medium (range) | 37 (0-117) |
| **Age categorized** |  |
| < 20 years old | 112,096 (13.66) |
| 21-39 years old | 346,013 (42.17) |
| 40-59 years old | 239,856 (29.23) |
| >60 years old | 122,563 (14.94) |
| **Sex (women)** | 413,997 (50.43) |
| **Social stratum** |  |
| 1 – 3 | 484,183 (93.85) |
| 4 – 6 | 31,703 (6.15) |
| **HBP** | 4,632 (22.68) |
| **Mellitus diabetes** | 23,568(3.73) |
| **Obesity** | 20,236 (3.21) |
| **Smoking** | 11,417 (1.81) |
| **Respiratory disease** | 9,230 (1.55) |
| **Cancer** | 4,386 (0.70) |
| **Autoimmune disease** | 165 (0.81) |
| **Cardiovascular disease** | 1,524 (0.24) |
| **Cerebrovascular disease** | 360 (0.06) |
| **Pregnancy** | 2,563 (0.41) |
| **HIV** | 1,744 (0.28) |
| **Comorbidities** | 58,738 (9.31) |
| **Severity of case** | 23,374 (2.4) |

\* The source of the data used for these analyses is SegCovid and SIVIGILA. Absolute and percentage values are presented. In addition, a categorization of the age variable is made.

#### Algorithm development and external validity

After multiple calibrations, we generated four risk categories according to the final ROC curves results. The first risk category was *very high priority*: comprised of cases aged 60 or over or with comorbidities. We obtained a sensitivity of 81.4%, 84.9% of specificity, and an area under curve (AUC) by 83% for this category. The second risk category of priority was *high priority*, comprised of men between 40 to 59 years old without comorbidities, and it obtained a sensitivity by 75.9%, 67.3% of specificity, and an AUC by 71%. The third risk category of priority comprises men under 40 years old from low SES or pregnant women, all of them without comorbidities. We obtained a sensitivity by 10.9%, 87.1% of specificity, and an AUC by 49% for this third category. The last risk category was the *low priority* category comprised of those who do not classify any of the three previous ones, such as women under 60 years old without pregnancy or men under 40 years of social stratum from 4 to 6. We obtained a sensitivity of 6.8%, 37.3 of specificity, and an AUC by 22% for this category. Lastly, we identified that our algorithm has 91% sensitivity for detecting severe COVID-19 cases when taking together the first two risk categories (*very high* and *high priority*). The distribution of cases by category can be seen in table 3, and operational characteristics for the different categories can be seen in table 4. The full algorithm is shown in figure 2. Appendix 1 includes a field manual based on the algorithm.

**Table 3.** Distribution of observations by risk category.

| **Risk category** | **Number of severe COVID-19 cases** | **Total observations classified by category** | **Percentage of all severe COVID-19 cases classified in each category** |
| --- | --- | --- | --- |
| Very high priority | 19,040 | 300,551 | 81% |
| High priority | 2,268 | 109,342 | 10% |
| Medium priority | 483 | 104,359 | 2% |
| Low priority | 1,583 | 457,807 | 7% |
| Total | **23,374** | **972,059** | **100%** |

**Table 4.** Operational characteristics for the risk categories.

| **Risk category** | **Severe or fatal COVID-19** |  |  |  |
| --- | --- | --- | --- | --- |
|  | *Sensitivity* | *Specificity* | *AUC* | *Std. Err.* |
| Very high priority | **81.46%** | 84.85% | 83% | 0.00 |
| High priority | **75.90%** | 67.32% | 71% | 0.00 |
| Medium priority | **10.91%** | 87.11% | 49% | 0.00 |
| Low priority | **6.82%** | 37.37% | 22% | 0.00 |

\* ROC curve analysis where the dependent variable was case severity (those who reported having entered intermediate, intensive care or being dead from COVID-19). The independent variable was the priority category *Very High*, *High*, *Medium* or, *Low*. *AUC*: area under curve and *Std. Err.*: standard error.

**Figure 2.**
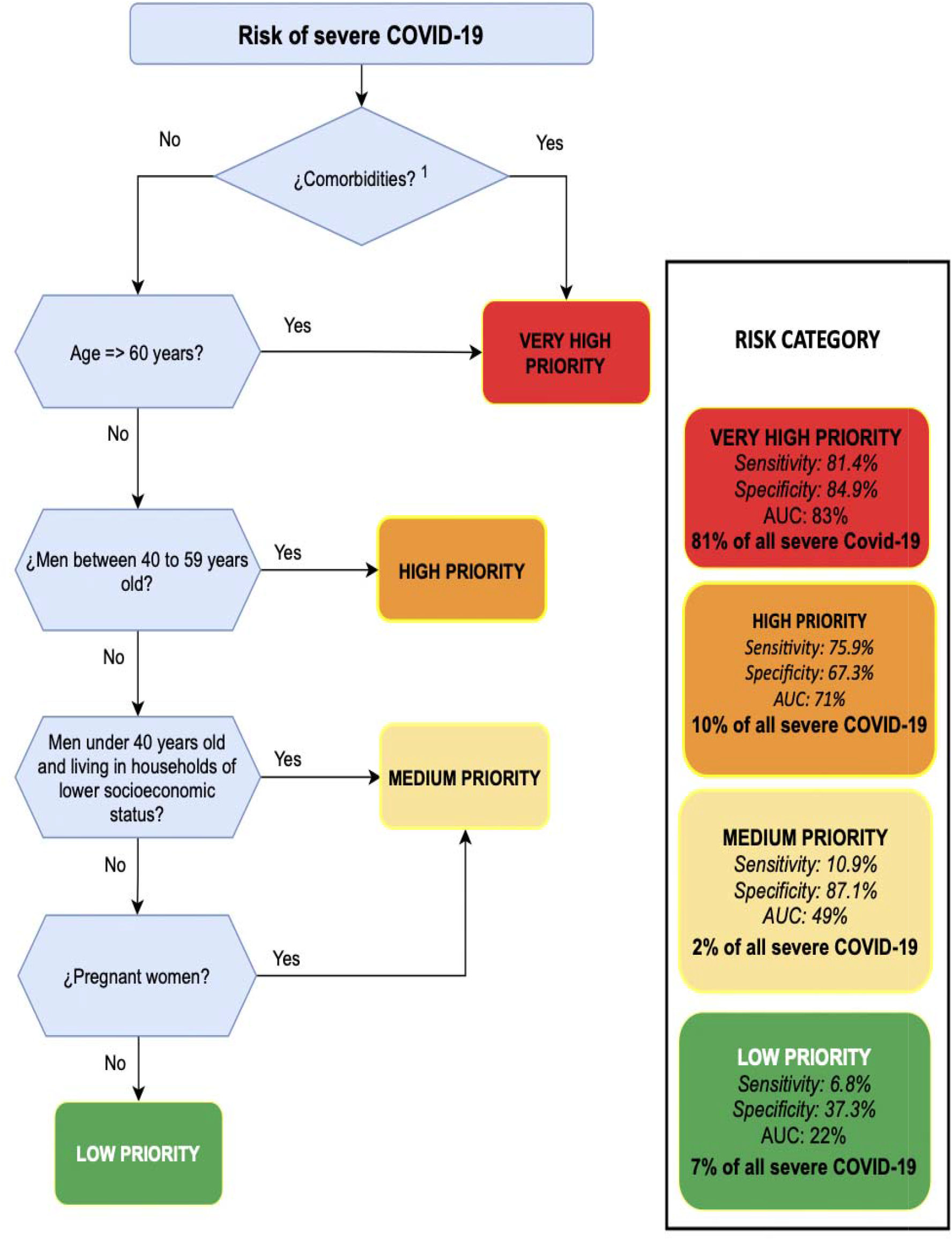
Risk monitoring algorithm. ^1^Cancer, HIV, cerebrovascular disease, high blood pressure(HBP), heart disease, immunosuppressed, cardiovascular disease, diabetes mellitus, smoking, obesity, and kidney disease

## DISCUSSION

In this study, we conducted a literature review and developed a prioritization tool to improve monitoring for COVID-19 cases. This tool is particularly relevant for contexts where public health teams have limited resources to conduct monitoring activities for COVID-19 patients and, more broadly, all types of public health measures during the pandemic.

In this study, we developed an algorithm with four different risk categories based on standard observable variables by public health teams. This algorithm allowed us to identify in the first two risk categories 91% of all patients reported to have severe COVID-19. This tool helps to focus limited resources on those who are more likely to experience severe COVID-19, especially when effective treatments, such as oxygen, corticoid therapy, or other strategies have been identified to reduce lethality associated to COVID-19 infection ^1,8^.

This algorithm is a well-suited tool for resource-constrained environments, but not necessarily limited to lower and middle-income countries (LMIC), as public health teams in higher-income countries have also found themselves overwhelmed with the size of the different pandemic waves.

This study has some limitations. First, we conducted a literature review that might have overlooked some important variables that can potentially be included. Second, this work relies on the accuracy of the two data systems used for this study. Other studies have shown that the Colombian registries for COVID-19 cases are of good quality, and there are no reasons to believe that a significant number of critical cases or deaths have been underreported ^2,9^.

This tool could be easily adapted to the context of other LMICs as data requirements are often available. This tool’s advantage is its low administrative cost, low organizational upfront investment, and flexibility to be improved over time as the demand for more effective monitoring changes. Implementing this tool will complement traditional public health strategies such as contact tracing, social distancing, lockdowns, etc.

Finally, implementation needs to be a priority in the planning process. This tool will require the development of monitoring systems that will provide benefits in future pandemics. When monitoring is outside the realm of public institutions and involves negotiations with the private sector for its implementation, this might lead to additional implementation barriers. Appropriate monitoring needs concrete interventions to make it effective, including the provision of oximeters, home-based health care services, and the implementation of call centers.

This study is a fundamental tool to improve public health teams’ responsiveness and efficiency to handle COIVD-19 cases, particularly during outbreaks in both LMIC and higher-income countries. Monitoring systems for COVID-19 patients is a critical long-term strategy, especially with limited access to vaccines and the possibility of these losing efficacy against new virus strains.

## Data Availability

The anonymized datasets analyzed during this study are available from Colombian Ministry of Health and Social Protection (MSPS). Data cannot be shared publicly because of privacy reasons as it contains individual-level information on demographics and health conditions. Researchers interested in obtaining these data can contact the Epidemiology and Demography Office at the MSPS for questions about data access requirements.

## Ethical Approval

This study was approved by the Institutional Review Board of the Johns Hopkins Bloomberg School of Public Health and deemed not human subjects research (IRB number: 14144).

## Acknowledgments

Many thanks to Yesika Hernandez for her support in designing the figures of appendix 1.

## Authors contributions

AVO conceived the methodological design, conceived the original approach, led the literature review and data analysis, and drafting the manuscript.

NG conducted the data analysis, contributed to the literature review, contributed to the methodological design, and drafting the manuscript.

YG conducted the data analysis, contributed to the literature review, contributed to the methodological design, and drafting the manuscript.

EPQ contributed to the methodological design and to drafting the manuscript.

JFN contributed to the original approach and contributed to drafting the manuscript. FR contributed to drafting the manuscript.

AJT obtained the funding, contributed to the methodological design and drafting the manuscript

---

**Appendix 1.** Risk monitoring field manual.

| STEP | Activity to be performed | Yes | NO |
| --- | --- | --- | --- |
| 1 | Is this person a confirmed case of covid-19? | Continue to STEP 2 | Re-evaluate the case |
| 2 | Identify if the person has any of the following disease(s) or condition(s):<br>• *History of current or past smoking (at least 100 cigarettes in lifetime).*<br>• *Cancer treatment*<br>• *HIV*<br>• *Cerebrovascular disease*<br>• *Immunosuppressed*<br>• *Cardiovascular disease*<br>• *Diabetes mellitus*<br>• *Obesity*<br>• *Chronic kidney disease (insufficiency)* | VERY HIGH PRIORITY | Continue to STEP 3 |
| 3 | Is the person over 60 years old? | VERY HIGH PRIORITY. | Continue to STEP 4 |
| 4 | Is the person a man? | Continue to STEP 5 | Continue to STEP 7 |
| 5 | Is the person a male between 40 and 59 years old? | HIGH PRIORITY. | Continue to STEP 6 |
| 6 | Is the person a male under 40 years of age living in a household of low socioeconomic status? | MEDIUM PRIORITY. | Continue to STEP 7 |
| 7 | Is the person a pregnant woman? | MEDIUM PRIORITY. | LOW PRIORITY. |

## References

1. The RECOVERY Collaborative Group. Dexamethasone in Hospitalized Patients with Covid-19 — Preliminary Report. New England Journal of Medicine 0, null (2020).

2. Vecino-Ortiz, A. I., Congote, J. V., Bedoya, S. Z. & Cucunuba, Z. M. Impact of contact tracing on COVID-19 mortality: An impact evaluation using surveillance data from Colombia. PLOS ONE 16, e0246987 (2021).

3. WHO Collaborating Centre for Infectious Disease Modelling. Report 4 - Severity of 2019 novel coronavirus (nCoV). http://www.imperial.ac.uk/medicine/departments/school-public-health/infectious-disease-epidemiology/mrc-global-infectious-disease-analysis/covid-19/report-4-severity-of-covid-19/ (2020).

4. Levin, A. T. et al.. Assessing the age specificity of infection fatality rates for COVID-19: systematic review, meta-analysis, and public policy implications. Eur J Epidemiol 35, 1123–1138 (2020).

5. Ministerio de Salud y Protección Social. SEGUIMIENTO EN SALUD A PERSONAS RESIDENTES EN COLOMBIA EN LA PANDEMIA DE COVID-19. https://www.minsalud.gov.co/RID/asif04-guia-seguimiento-casos-covid19-1.0.pdf (2020).

6. Vecino Ortiz, A. I. & Arroyo-Ariza, D. F. A tax on Sugar Sweetened Beverages in Colombia: Estimating the impact on overweight prevalence and revenue generation. Submitted (2016).

7. Departamento Administrativo Nacional de Estadística (DANE). Estratificación socioeconómica. https://www.dane.gov.co/index.php/servicios-al-ciudadano/servicios-informacion/estratificacion-socioeconomica.

8. University of Oxford. Randomised Evaluation of COVID-19 Therapy. https://clinicaltrials.gov/ct2/show/NCT04381936 (2021).

9. Ministerio de Salud y Protección Social. EXCESO DE MORTALIDAD EN COLOMBIA 2020. https://www.minsalud.gov.co/sites/rid/Lists/BibliotecaDigital/RIDE/VS/ED/VSP/estimacion-exceso-mortalidad-Colombia-2020.pdf (2020).

10. Lighter, J. et al.. Obesity in Patients Younger Than 60 Years Is a Risk Factor for COVID-19 Hospital Admission. Clin Infect Dis 71, 896–897 (2020).

11. Romero Starke, K. et al. The Age-Related Risk of Severe Outcomes Due to COVID-19 Infection: A Rapid Review, Meta-Analysis, and Meta-Regression. International Journal of Environmental Research and Public Health 17, 5974 (2020).

12. Li, X. et al.. Risk factors for severity and mortality in adult COVID-19 inpatients in Wuhan. J Allergy Clin Immunol 146, 110–118 (2020).

13. Pranata, R., Huang, I., Lim, M. A., Wahjoepramono, E. J. & July, J. Impact of cerebrovascular and cardiovascular diseases on mortality and severity of COVID-19– systematic review, meta-analysis, and meta-regression. Journal of Stroke and Cerebrovascular Diseases 29, 104949 (2020).

14. Huang, Y. et al.. Obesity in patients with COVID-19: a systematic review and meta-analysis. Metabolism 113, 154378 (2020).

15. Popkin, B. M. et al.. Individuals with obesity and COVID-19: A global perspective on the epidemiology and biological relationships. Obesity Reviews 21, e13128 (2020).

16. Kumar, A. et al.. Is diabetes mellitus associated with mortality and severity of COVID-19? A meta-analysis. Diabetes & Metabolic Syndrome: Clinical Research & Reviews 14, 535–545 (2020).

17. Public Health England. COVID-19: review of disparities in risks and outcomes. https://assets.publishing.service.gov.uk/government/uploads/system/uploads/attachment_data/file/908434/Disparities_in_the_risk_and_outcomes_of_COVID_August_2020_update.pdf (2020).

18. Alqahtani, J. S. et al.. Prevalence, Severity and Mortality associated with COPD and Smoking in patients with COVID-19: A Rapid Systematic Review and Meta-Analysis. PLOS ONE 15, e0233147 (2020).

19. Ellington, S. Characteristics of Women of Reproductive Age with Laboratory-Confirmed SARS-CoV-2 Infection by Pregnancy Status — United States, January 22 –7 June, 2020. MMWR Morb Mortal Wkly Rep 69, (2020).

20. Fernández-Niño, J. A., Guerra-Gómez, J. A. & Idrovo, A. J. Multimorbidity patterns among COVID-19 deaths: proposal for the construction of etiological models. Rev Panam Salud Publica 44, (2020).

21. UK Office for National Statistics. Coronavirus (COVID-19) related deaths by occupation, England and Wales: deaths registered up to and including April 20 2020. https://www.ons.gov.uk/peoplepopulationandcommunity/healthandsocialcare/causesofdeath/bulletins/coronaviruscovid19relateddeathsbyoccupationenglandandwales/deathsregistereduptoandincluding20april2020.

